# A Preliminary Retrospective and Prospective Cohort Study on a Traditional Chinese Long-term Extreme Fasting

**DOI:** 10.1101/2020.03.14.20036111

**Authors:** Chao Wang, Ligang Ming, Lijun Jia, Qi Wang, Tingting Cao, Liping Wang, Zijing Zhou, Dan Tong, Wei Li, Yiqing Wu, Hong Ding, Di Liu, Minghui Zhang

**Affiliations:** School of Medicine, Tsinghua University, Beijing, China; Cancer Institute of Longhua Hospital, Shanghai University of Traditional Chinese Medicine, Shanghai, China; Nanshan Branch of Qilu Hospital, Yantai, China; Institute of Microbiology, Chinese Academy of Sciences, Beijing, China; Institute of Virology, Chinese Academy of Sciences, Wuhan, China; Institute of Physical Science and Information Technology, Anhui University, Hefei, China; The 8th Medical Center of Chinese PLA General Hospital, Beijing, China

**Keywords:** Fasting, metabolic disorder, hypertension, hyperlipidemia, fatty liver

## Abstract

**Background:** Fasting has long been a ritual or practice in varied religions, and recently, has been noticed to reduce the risk factors of metabolic diseases. In China, varied populations performed a traditional Taoism fasting practice, which lasted for 21-day with <5% calorie intake. However, the safety and applicability of this procedure haven not been investigated.

**Methods:** A total of 144 volunteered participants in six camps following the 21-day fasting (with <5% of normal diet) were investigated. 124 were examined for physical biomarkers and 53 of which also had biochemical markers. Another open label, non-comparative, phase 1/2 prospective cohort study enrolling 20 participants with metabolic diseases was also performed. The physical indices and biochemical biomarkers were collected at varied point of the fasting procedure. Statistical comparison and metagenomic analysis were performed. This study was registered in ClinicalTrials.gov (NCT03193177).

**Findings:** Our preliminary retrospective cohort study showed that no severe adverse event (grade 3 or above) was reported, and all biomarkers fluctuated within the safe ranges, except for the urea acid. The 21-day fasting could significantly reduce BMI and blood pressures. The prospective cohort study of the metabolic diseased participants showed a significant reduction of BMI (3.3±1.0) and systolic blood pressure (28.7±17.8 mmHg) after the fasting procedure. The data also presented significant ameliorations on overweight (16/16), hypertension (11/11) and fatty liver (9/9).

**Interpretation:** The 21-day fasting appeared safe and feasible for both healthy and unhealthy people. It could ameliorate the risk factors associated with hypertension and hyperlipidemia.

**Funding:** This work was supported by National Natural Science Foundation of China and China Overseas-Educated Scholars Development Foundation.

**Research in context:** *Evidence before this study:* Fasting has long been a ritual or practice in varied religions. In modern science, it has been noticed that fasting or calorie-restricted diets could benefit for the prevention or treatment of metabolic disorder-associated diseases. In China, the fasting practice, called “Bigu” (literally: avoiding grains), is believed to be capable of prolonging life in Taoism and was also used for medical cure. Compared to the reported fasting practices, the Bigu regimen is a more restricted abstinence, in which the practicers usually experience a continuous 21-day practice with an extremely low-calorie intake (<5% of normal diet). In a rough estimation, there are dozens of Bigu practice camps and over ten thousand practicers per year in China. However, nearly all Bigu camps followed the traditional Taoist procedures but lacked medical and scientific evaluation, which made those practices either mysterious or superstitious to the public.

*Added value of this study:* Our data showed that no severe adverse event was reported during the 21-day fasting procedure, and all biomarkers fluctuated within the safe ranges, except for the urea acid. The 21-day fasting could significantly reduce BMI and blood pressures. The data also presented significant ameliorations on overweight, hypertension and fatty liver. This 21-day fasting appeared safe and feasible for both healthy and unhealthy people. It could ameliorate the risk factors associated with hypertension and hyperlipidemia.

*Implications of all the available evidence:* This preliminary cohort study showed that the long-term extreme fasting was safety to most people and exhibited promising therapeutic effects to hypertension, hyperlipidemia and fatty liver. However, a large cohort study of health-improving effects by long-term extreme fasting is needed.

## Introduction

Fasting features abstinence or reduction of food, drink or both in a period of time, and is normally acted as ritual in many religions. Muslims incorporate periods of fasting into their rituals from dawn until dusk during the month of Ramadan; Hindus fast on certain days of a month; while some Christian denominations follow the acceptable fast according to Bible. In China, the fasting practice, called “Bigu” (literally: avoiding grains), are believed to be capable of prolonging life in Taoism and was also used for medical cure. In modern science, it has been noticed that fasting or calorie-restricted diets could benefit for the prevention or treatment of cardiovascular diseases(*1, 2*), autoimmune diseases(*3*), cancers(*2, 4*), diabetes(*5-7*), and also proved their applicability in tumor suppression(*8-11*), life extension(*12*), neural repair(*13*) and autoimmune remission(*14*) in mice. Particularly, some clinical trials focusing on short time (less than 7 consecutive days, 36-56% of normal diet) fasting-mimicking diet(*2, 15*) or prolonged low intensity (6 months, approximately 75% of normal diet) calorie restriction diet(*1, 6, 7*) have been carried out, and the safety and capability to improve health were well examined.

Compared to the above fasting practices (fasting-mimicking diet and calorie restriction diet), the Bigu regimen is a more restricted abstinence, in which the participants usually experience a continuous 21-day practice with an extremely low-calorie intake (<5% of normal diet) under the supervision of Bigu Masters. In a rough estimation, there are dozens of Bigu practice camps and over ten thousand participants per year in China and become popular in both aging population and middle-aged high-income population. Of special note, nearly all Bigu camps followed the traditional Taoist procedures and lacked medical and scientific evaluation, which made those practices either mysterious or superstitious to the public. Hence, whether the long-time extreme fasting process could result in safe and effective outcome requires scientific investigation.

In this study, we investigated 144 volunteer Bigu participants in six camps, which followed the 21-day regimen (with <5% of normal diet). The first 124 volunteers were collected for physical biomarkers and 53 of which have both physical and biochemical markers during the entire procedure. The data showed that the biomarkers return to normal ranges in about 10 days post the end of fasting, and the participant showed no severe adverse effect. Interestingly, we noticed that some participants with cardiovascular diseases had the previous aberrant physical markers back to the healthy levels after the whole fasting procedure. Of special interest, we prospectively followed 20 participants with aberrant biomarkers of cardiovascular diseases, and tracked their physical indications at pre-fasting, fasting and post-fasting. The results showed that 16 participants with higher BMI, 11 with hypertension and 9 with fatty liver got a significant amelioration, indicative of its potential therapeutic effects. In general, this preliminary cohort study showed that the long-term extreme fasting was safe to both healthy and unhealthy people and exhibited promising therapeutic effects.

## Materials and Methods

### Long-term extreme fasting (Bigu regimen) Procedures

All subjects receiving Bigu regimen should stay together for at least the first three days with the supervision by an expert. Exercises and meditation should be performed twice a day (usually once in the morning and once in the afternoon). The whole process of Bigu regimen was divided into two sections. For the first phase, participants should be maintained in a water-only or extremely low-calorie intake diet for 21 days under the instruction of an experienced supervisor. The source of calorie was mainly from brown sugar water and honey water (∼141.8kJ), each of which should be taken less than 1000mL per day. Participants who suffered from severe starvation could have 4g of dates, 3g of peanuts and 2g of walnuts (∼138.1kJ). Comparison of macronutrient and micronutrient intake between this fasting procedure and normal diet was shown as Table 2. For the second phase, these participants resumed taking foods incrementally for the first seven days (from day 22 to day 28). A detailed recommended diet composition during the 7-day resuming period was listed as Table 3. Participants could not take normal diets until finishing the second phase in order to avoid accidental damage to the gastrointestinal tract.

**Table 1.** Patient demographics, baseline characteristics and biomarker changes after bigu regimen.

|  | A basic survey<br>(n=124) | A detailed Analysis<br>(n=53) | A prospective trial<br>(n=20) |
| --- | --- | --- | --- |
| Median age (years) | 47±10 | 50±10 | 60±13 |
| ≥60 years | 14(11%) | 9(17%) | 10(50%) |
| Sex |  |  |  |
| Female | 76(61%) | 34(64%) | 13(63%) |
| Male | 48(39%) | 19(36%) | 7(37%) |
| Weight (kg) |  |  |  |
| Baseline | 69±15 | 73±16 | 74±21 |
| Day 21 of bigu | 61±13 | 62±13 | 65±19 |
| BMI |  |  |  |
| Baseline | 24.7±4.5 | 25.0±4.1 | 26.5±5.1 |
| Day 21 of bigu | 21.8±4.0 | 21.7±3.5 | 23.4±4.6 |
| SBP (mmHg) |  |  |  |
| Baseline | 126±17 | 127±19 | 143±25 |
| Day 21 of bigu | 114±12 | 110±12 | 117±21 |
| DBP (mmHg) |  |  |  |
| Baseline | 82±14 | 83±15 | 79±12 |
| Day 21 of bigu | 74±9 | 71±10 | 76±11 |
| Triglycerides (mmol/L) |  |  |  |
| Baseline | nd | 1.8±1.4 | 1.6±0.8 |
| Day 21 of bigu | nd | 1.3±0.5 | 1.4±0.3 |
| Total cholesterol(mmol/L) |  |  |  |
| Baseline | nd | 5.9±1.3 | 5.5±1.1 |
| Day 21 of bigu | nd | 5.0±1.2 | 5.7±0.9 |
| LDL (mmol/L) |  |  |  |
| Baseline | nd | 4.1±1.0 | 3.3±0.7 |
| Day 21 of bigu | nd | 3.4±0.9 | 3.5±0.6 |
| HDL (mmol/L) |  |  |  |
| Baseline | nd | 1.4±0.4 | 1.3±0.2 |
| Day 21 of bigu | nd | 1.2±0.4 | 1.1±0.1 |
| Glucose (mmol/L) |  |  |  |
| Baseline | nd | 5.3±2.3 | 6.1±3.2 |
| Day 21 of bigu | nd | 4.9±2.0 | 4.8±1.2 |
| Uric acid (mmol/L) |  |  |  |
| Baseline | nd | 403.0±113.4 | 326.1±114.0 |
| Day 21 of bigu | nd | 462.6±229.8 | 636.6±202.4 |
| ALT (U/L) |  |  |  |
| Baseline | nd | 21.0±12.3 | 21.9±14.8 |
| Day 21 of bigu | nd | 25.1±18.9 | 34.6±43.7 |
| AST (U/L) |  |  |  |
| Baseline | nd | 25.9±14.1 | 22.8±9.1 |
| Day 21 of bigu | nd | 31.7±18.2 | 37.7±29.0 |
| BUN (mmol/L) |  |  |  |
| Baseline | nd | 5.7±1.4 | 6.5±1.3 |
| Day 21 of bigu | nd | 3.6±1.2 | 4.7±1.2 |

**Table 2.** Nutrition composition during bigu regimen.

| Categories | Bigu | Normal | Ratio(%) |
| --- | --- | --- | --- |
| Calories(KJ) | 283.58 | 8263.40 | 3.43 |
| Total Fat(g) | 2.12 | 65.84 | 3.21 |
| Carbohydrate(g) | 10.21 | 320.94 | 3.18 |
| Dietary Fiber(g) | 0.60 | 25.00 | 2.40 |
| Protein(g) | 0.85 | 55.00 | 1.55 |
| Vitamin A(μg) | 0.28 | 750.00 | 0.04 |
| Vitamin C(mg) | 1.00 | 100.00 | 1.00 |
| Vitamin E(mg) | 1.28 | 14.00 | 9.17 |
| Na(mg) | 10.41 | 1450.00 | 0.72 |
| Ca(mg) | 9.26 | 900.00 | 1.03 |
| Fe(mg) | 0.38 | 14.00 | 2.70 |

**Table 3.** Nutrition composition during the 10-day recovery diet.

| From bigu | Day 22 | Day 23 | Day 24 | Day 25 | Day 26 | Day 27 | Day 28 | Day 29-31 |
| --- | --- | --- | --- | --- | --- | --- | --- | --- |
| From resuming diet | Day 1 | Day 2 | Day 3 | Day 4 | Day 5 | Day 6 | Day 7 | Day 8 |
| **Calories(kcal)** | 347.44 | 694.88 | 1529.75 | 2545.63 | 3610.21 | 5409.87 | 6342.90 | 8325.99 |
| **Total Fat(g)** | 1.13 | 2.26 | 4.68 | 9.16 | 10.32 | 27.11 | 28.21 | 41.30 |
| **Cabohydrate(g)** | 14.47 | 28.94 | 61.27 | 109.39 | 165.41 | 228.59 | 274.29 | 342.04 |
| **Protein(g)** | 3.27 | 6.54 | 17.16 | 22.86 | 29.03 | 38.75 | 45.75 | 69.01 |

### Patient cohorts

This study (ClinicalTrials.gov; NCT03193177) contained three progressive sections (Table 1). We firstly took a basic survey on 124 people who voluntarily performed the Bigu regimen for 21 days. Of these 124 subjects, 53 participants were completely followed before, during and after Bigu regimen when their weight, blood pressure and blood biochemistry biomarkers were examined. According to these survey results, we prospectively followed 20 participants with aberrant biomarkers of cardiovascular diseases, and tracked their physical indications at pre-fasting, fasting and post-fasting. This study was conducted in Nanshan Branch of Qilu Hospital from June to September in the year 2017 and 20 subjects were voluntarily enrolled into this observational study. Of these 20 participants, 18 had succeeded in accomplishing the entire period of fasting, 1 withdrew on day 8 because of scheduling conflicts and 1 withdrew on day 14 due to the noncompliance of Bigu regimen. All participants have signed voluntary informed consent for collection and analysis of blood and fecal microbiome samples under Institutional Review Board (IRB)–approved protocols (THUMED-BG-170612). All clinical examination and evaluation was carried out independently and recorded instantaneously.

### Statistical analysis

After verify normally distribution of the data, comparison between two time points (e.g. day 0 versus day 21) was performed using paired two-tailed Student’s *t* tests. When the data does not follow normal distribution, comparison was carried out using Wilcoxon signed-rank test. *P* values <0.05 were considered significant. *** means *P*<0.001, ** means 0.001<*P*<0.01 and * means 0.01<*P*<0.05.

## Results

### A basic survey of physical markers follows 124 bigu participants

From March 2016 to Oct 2017, we followed 124 people who carried out 21-day Bigu regimen and recorded their physical markers just pre-fasting (day 0), mid-fasting (day 10), end-of-fasting (day 21) and 10-day post fasting (day 31) (Figure 1A). Of these 124 participants, 48 (39%) were male and 76 (61%) were female. During fasting, the participants were asked to report any physical discomfort in time. The most common self-reported grade 1 symptom is fatigue, while dry mouth, tingling and nausea were also listed. Weakness, vomiting, asthma and fever has been reported occasionally as the grade 2 symptoms. No severe adverse event was reported as grade 3 and above (Figure 1B). We investigated the normal activities of each participant by using a detector MI Band worn on the wrist, to record walking distance, heart rates, sleeping time and consumed energy. The data presented no significant difference during fasting compared to those at day 0 (Figure 1C), indicating those participants kept daily activities during the procedure. These data suggested that the long-term extreme (21-day, <5% of normal diet) fasting procedure causes no adverse events beyond grade 2 and is likely be safe to most normal people.

**Figure 1.**
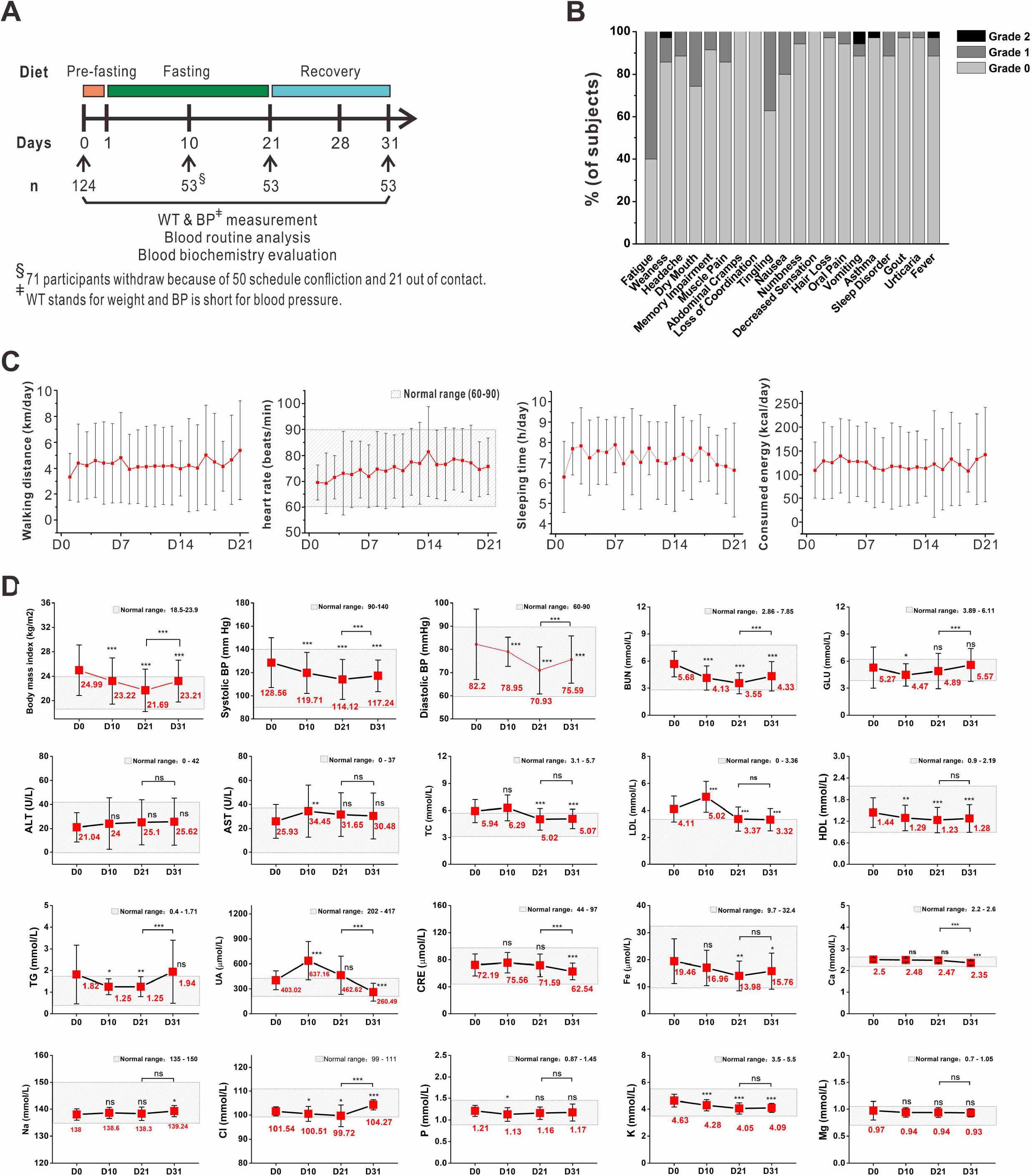
A preliminary survey of physical biomarkers in 53 subjects during 21-day fasting procedure. **A**. Schema of fasting procedure in this survey. **B**. Self-reported adverse effects were recorded according to CTCAE, where 0=No symptoms, 1=Mild, 2=Moderate, 3=Severe, 4=Life threatening, 5=Death. **C**. Daily activities were recorded using Mi Band. **D**. Basic physical biomarkers were examined during the entire fasting procedure.

### A detailed analysis on physical biomarkers of 53 participants

We further reviewed the data of all 124 participants, 53 of which had serial physical indices. Among the 20 examined indices, the BMI, diastolic blood pressure and systolic blood pressure decreased significantly in participants during fasting, compared to day 0 (Figure 1D). Meanwhile, some metabolic biomarkers, including blood urea nitrogen (BUN), total cholesterol (TC), triglyceride (TG), low density lipoprotein (LDL), creatinine (CRE) and high density lipoprotein (HDL) also declined during this procedure (Figure 1D). No microelements fell below normal levels, though Fe, Ca, Cl and P showed slightly decreases (Figure 1D). Unexpectedly, the fasting glucose was not detected decrease on day 21 (end-of-fasting), ruling out hypoglycemia during this period (Figure 1D). These data indicated that the long-term extreme fasting did not cause severe deviations out of the normal ranges of those physical indices, though as expected, some biomarkers (e.g. uric acid, triglyceride) had a significant fluctuation. The results also presented that the fasting procedure could decrease the blood pressure and blood cholesterol associated biomarkers, while kept blood glucose balanced, implying its potential therapeutic effects on hypertension and hyperlipidemia.

### A prospective study on 20 participants with hypertension or hyperlipidemia

In order to examine whether this long-term extreme fasting would benefit those people with hypertension and/or hyperlipidemia, we prospectively recruited 20 unhealthy people (in one Bigu camp) and detected their dynamic changes of metabolic physical biomarkers, from June 18^th^ to Sep 8^th^ in the year 2017. Among the 20 participants, 7 (35%) were male and 13 (65%) were female. Eighteen of 20 participants had succeeded in accomplishing the entire period of fasting, 1 withdrew on day 8 because of scheduling conflicts and 1 withdrew on day 14 due to the noncompliance of Bigu regimen (Figure 2A). Of note, we kept collecting data from those 18 participants until day 51 (30-day post resuming). Before fasting (examined on day 0), 16 participants were overweight, 11 suffered from hypertension, 9 were hyperlipidemia, 3 were hyperglycemia, and 9 had fatty liver (Table 4).

**Table 4.** Physical examination of 20 participants before fasting procedure (day 0). ^§^ indicates the withdrawn case from this clinical trial.

|  | Overweight | Hypertension | Hyperlipidemia | Hyperglycemia | Fatty Liver |
| --- | --- | --- | --- | --- | --- |
| Case 1 | ✓ | ✓ | ✓ | ✓ | none |
| Case 2 | ✓ |  |  |  | none |
| Case 3 | ✓ |  |  |  | none |
| Case 4 | ✓ | ✓ | ✓ |  | mi to mo |
| Case 5 | ✓ | ✓ |  |  | none |
| Case 6 | ✓ | ✓ | ✓ |  | none |
| Case 7 | ✓ |  | ✓ |  | none |
| Case 8 |  | ✓ |  |  | none |
| Case 9 | ✓ |  | ✓ |  | moderate |
| Case 10 | ✓ | ✓ |  |  | severe |
| Case 11<sup>§</sup> | ✓ |  |  |  | moderate |
| Case 12 | ✓ |  |  |  | none |
| Case 13 | ✓ | ✓ | ✓ |  | mi to mo |
| Case 14 |  |  | ✓ | ✓ | none |
| Case 15 | ✓ | ✓ | ✓ |  | none |
| Case 16 | ✓ | ✓ |  |  | mild |
| Case 17 | ✓ | ✓ |  |  | severe |
| Case 18<sup>§</sup> |  |  | ✓ |  | none |
| Case 19 | ✓ | ✓ |  | ✓ | moderate |
| Case 20 |  |  |  |  | mi to mo |
| Total | 16 | 11 | 9 | 3 | 9 |

**Figure 2.**
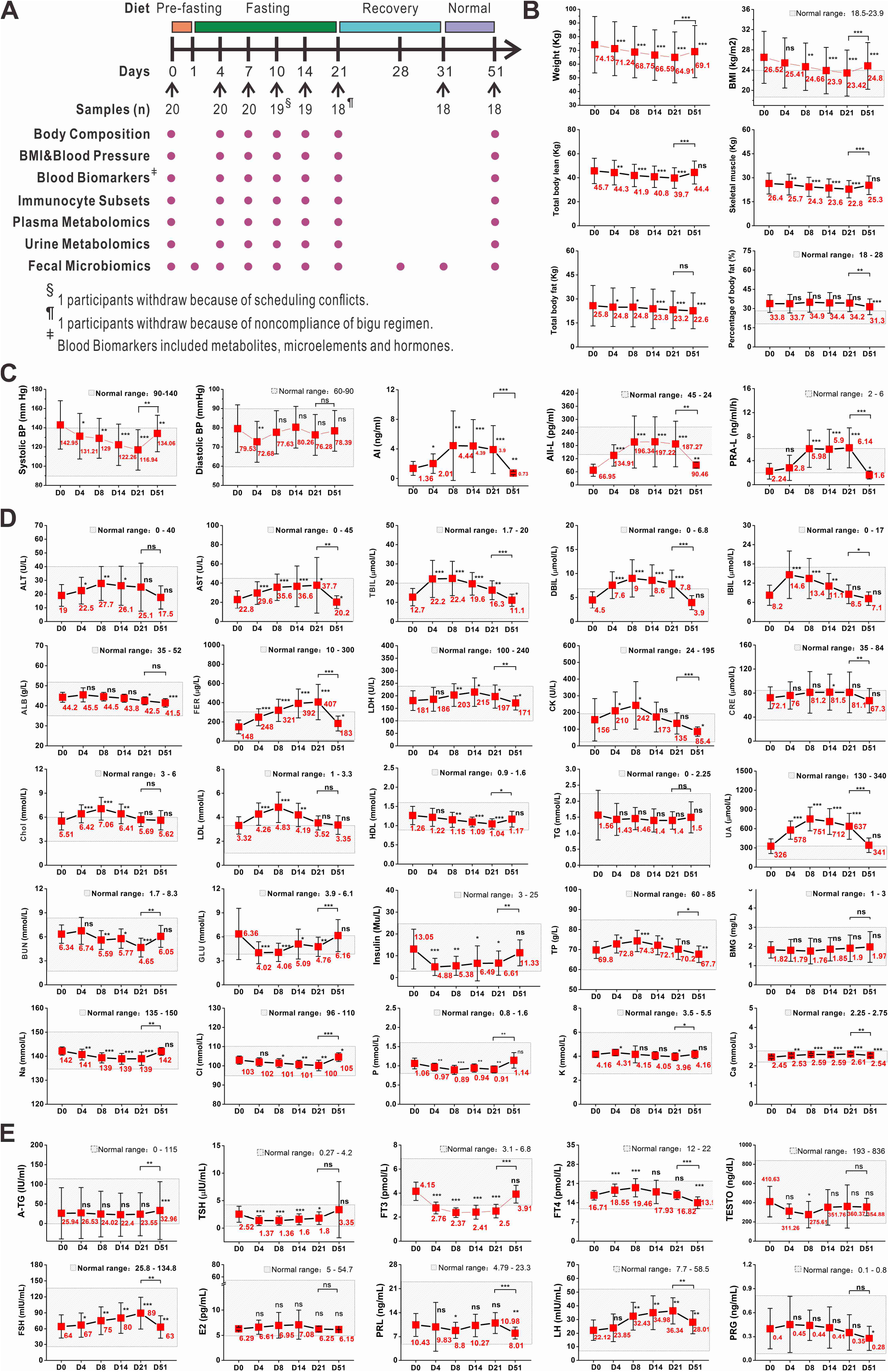
A deep investigation of physical biomarkers in 20 subjects with metabolic diseases. **A**. Schema of fasting procedure in the clinical trial. **B-E**. Changes of non-invasive physical indexes **(B)**, hypertension associated biomarkers **(C)**, biochemical biomarkers **(D)** and hormone levels **(E)** were evaluated during the fasting procedure.

We following carried out physical examination and biomarker detection on day 0, day 4, day 7, day 14, day 21 and day 51 (Figure 2A). The data showed that the 21-day fasting resulted in a significant reduction by 9.2 ±3.2 kg (P < 0.001) of body weight, accordance with a reduction by 3.3 ±1.0 of BMI (Figure 2B). Systolic blood pressure was reduced by 28.7± 17.8 mmHg, whereas diastolic blood pressure did not reach a significant change. Levels of three markers associated with hypertension including rennin, angiotensin I and angiotensin II were detected significant increases (P < 0.001, Figure 2C), indicating a feedback to elevate blood pressure through the rennin-angiotensin system. We also found that both total body lean and total body fat were significantly decreased during fasting process (Figure 2B), indicating the consumption of both muscles and fats for energy production.

Since most of the participants recruited suffered from metabolic diseases, we followed their metabolism associated biomarkers during the fasting procedure (Figure 2D). In the whole period, the concentrations of Na, Cl, P, K and Ca were not found to deviate beyond their normal ranges, while they still experienced fluctuations. The data showed that the levels of alanine transaminase, aspartate transaminase, total bilirubin, indirect bilirubin, direct bilirubin, ferritin, lactate dehydrogenase and creatine kinase were increased significantly. As glucose metabolic pathway was inhibited due to the lack of carbohydrate intake, levels of total cholesterol and low density lipoprotein, as well as total protein were elevated transiently during this period, but the level of high density lipoprotein was found to be decreased. The level of uric acid was found to be significantly increased after starting fasting, which did not return to the normal level until resuming normal diet (Figure 2D). The glucose level was decreased during the first seven days of fasting, while it started to increase on day 14, which was beyond our expectations. However, the insulin level started to decrease just after starting fasting and remained the lower level during the whole procedure.

As hormone plays an important role in regulating physical activities, dynamics of the levels of thyroid hormones and sex hormones were recorded at indicated time points (Figure 2E). For thyroid hormones, both TSH and serum free T3 levels were decreased significantly during fasting, the level of T4 was increased transiently on day 14, but ATG level did not show an obvious change. But among sex hormones including luteinizing hormone, follicle-stimulating hormone, prolactin, estradiol, progesterone and testosterone, only the levels of follicle-stimulating hormone and luteinizing hormone in women significantly increased during fasting procedure and maintained at the higher levels even on day 51.

Then we evaluate the effects of the 21-day fasting on metabolic diseases by assessing the changes of biomarkers and imaging examinations (Table 5). We examined the indices of all participants on day 21. The data showed that among the 12 participants with higher BMI (>23.9), 6 were reduced to the normal range, and the other 6 had a significant decrease (P<0.001). Of 11 participants suffering from hypertension, 9 decreased to the normal range, and the remaining 2 also had a remarkable decrease. Besides, the long-time extreme fasting were proved to be effective on fatty liver. We found that in 9 participants with fatty liver, 7 were ameliorated, including 2 from severe to moderate or mild, 4 from moderate to mild, and 1 from mild to non-fatty liver (Figure S1A-I). Conclusively, these data suggested that bigu regimen could benefit the unhealthy people in the high BMI, hypertension and fatty liver.

**Table 5.** Changes of individual biomarkers and imaging examinations during fasting procedure.

|  | Body Mass Index |  |  |  |  |  | Systolic Blood Pressure (mmHg) |  |  |  |  |  | Diastolic Blood Pressure (mmHg) |  |  |  |  |  | Glucose (mmol/L) |  |  |  |  |  |
| --- | --- | --- | --- | --- | --- | --- | --- | --- | --- | --- | --- | --- | --- | --- | --- | --- | --- | --- | --- | --- | --- | --- | --- | --- |
|  | Day0 | Day4 | Day8 | Day14 | Day21 | Day51 | Day0 | Day4 | Day8 | Day14 | Day21 | Day51 | Day0 | Day4 | Day8 | Day14 | Day21 | Day51 | Day0 | Day4 | Day8 | Day14 | Day21 | Day51 |
| Case 1 | 26.0 | 24.3 | 23.7 | 22.9 | 22.5 | 23.2 | 162 | 156 | 133 | 137 | 138 | 204 | 77 | 73 | 69 | 78 | 87 | 100 | 9.95 | 4.45 | 3.93 | 5.64 | 7.09 | 8.75 |
| Case 2 | 23.9 | 23.4 | 22.4 | 21.7 | 21.0 | 21.9 | 120 | 108 | 114 | 99 | 92 | 121 | 69 | 55 | 64 | 64 | 64 | 75 | 5.41 | 3.44 | 3.67 | 4.08 | 4.09 | 5.36 |
| Case 3 | 24.0 | 23.2 | 23.1 | 22.3 | 21.8 | 23.6 | 131 | 129 | 139 | 134 | 124 | 141 | 73 | 71 | 79 | 86 | 80 | 81 | 4.81 | 3.57 | 4.04 | 3.99 | 3.74 | 4.70 |
| Case 4 | 29.6 | 28.0 | 27.4 | 27.1 | 25.7 | 27.9 | 141 | 166 | 151 | 127 | 116 | 131 | 86 | 93 | 101 | 89 | 78 | 79 | 5.18 | 3.52 | 3.65 | 4.44 | 4.44 | 5.11 |
| Case 5 | 23.9 | 23.5 | 23.1 | 22.1 | 21.9 | 23.3 | 143 | 107 | 99 | 92 | 96 | 134 | 96 | 69 | 65 | 68 | 67 | 88 | 5.41 | 3.89 | 3.53 | 4.84 | 4.21 | 5.21 |
| Case 6 | 25.8 | 24.8 | 24.1 | 23.3 | 22.7 | 23.9 | 185 | 150 | 138 | 138 | 131 | 179 | 104 | 83 | 90 | 85 | 86 | 103 | 5.00 | 3.09 | 3.30 | 3.07 | 4.16 | 4.98 |
| Case 7 | 27.1 | 25.8 | 24.6 | 24.0 | 23.3 | 24.6 | 139 | 151 | 126 | 94 | 95 | 111 | 83 | 90 | 92 | 71 | 68 | 67 | 4.74 | 4.58 | 6.33 | 6.68 | 6.06 | 5.35 |
| Case 8 | 19.9 | 18.8 | 17.9 | 17.3 | 16.5 | 18.2 | 143 | 136 | 123 | 123 | 103 | 131 | 80 | 76 | 80 | 77 | 65 | 75 | 5.04 | 2.66 | 2.74 | 3.02 | 3.48 | 4.83 |
| Case 9 | 31.2 | 31.1 | 29.3 | 27.8 | 26.7 | 27.4 | 128 | 128 | 121 | 123 | 117 | 120 | 82 | 72 | 83 | 88 | 79 | 96 | 5.71 | 5.79 | 6.22 | 7.12 | 6.57 | 7.06 |
| Case 10 | 44.3 | 42.4 | 40.7 | 39.6 | 38.7 | 39.3 | 159 | 132 | 142 | 137 | 118 | 141 | 78 | 73 | 91 | 90 | 82 | 88 | 4.68 | 3.53 | 4.50 | 5.48 | 5.29 | 5.60 |
| Case 11 | 29.6 | 28.5 | wd | wd | wd | wd | 116 | 114 | wd | wd | wd | wd | 70 | 62 | wd | wd | wd | wd | 5.36 | 3.50 | wd | wd | wd | wd |
| Case 12 | 23.9 | 22.4 | 21.7 | 20.8 | 19.8 | wd | 117 | 108 | 106 | 110 | 103 | wd | 66 | 66 | 62 | 72 | 62 | wd | 5.08 | 3.20 | 4.10 | 4.20 | 4.66 | wd |
| Case 13 | 28.6 | 27.4 | 27.1 | 26.7 | 25.2 | 27.4 | 167 | 162 | 160 | 148 | 150 | 137 | 80 | 76 | 76 | 85 | 92 | 74 | 5.58 | 3.88 | 3.50 | 4.86 | 4.51 | 6.26 |
| Case 14 | 22.6 | 21.8 | 21.4 | 20.7 | wd | wd | 94 | 82 | 111 | 96 | wd | wd | 63 | 60 | 71 | 72 | wd | wd | 18.08 | 7.74 | 6.37 | 11.29 | wd | wd |
| Case 15 | 24.9 | 23.4 | 22.7 | 22.5 | 21.4 | 21.2 | 176 | 149 | 118 | 130 | 125 | 143 | 103 | 89 | 84 | 107 | 93 | 98 | 4.65 | 3.04 | 3.49 | 5.19 | 3.48 | 5.52 |
| Case 16 | 28.3 | 27.0 | 26.1 | 24.9 | 24.5 | 27.3 | 167 | 145 | 175 | 148 | 133 | 158 | 76 | 74 | 83 | 89 | 81 | 71 | 5.70 | 3.53 | 2.27 | 4.96 | 3.65 | 6.44 |
| Case 17 | 27.9 | 26.6 | 25.9 | 25.2 | 25.1 | 26.6 | 154 | 147 | 151 | 164 | 164 | 160 | 69 | 71 | 78 | 80 | 83 | 70 | 3.50 | 3.73 | 4.03 | 4.63 | 5.06 | 6.19 |
| Case 18 | 22.0 | 21.5 | 21.3 | 20.6 | 21.0 | 21.1 | 123 | 118 | 110 | 101 | 96 | 130 | 63 | 62 | 60 | 62 | 59 | 68 | 3.27 | 3.73 | 3.92 | 4.15 | 4.37 | 5.47 |
| Case 19 | 26.4 | 24.8 | 24.1 | 23.4 | 23.8 | 23.5 | 166 | 127 | 133 | 128 | 116 | 192 | 93 | 72 | 78 | 85 | 81 | 103 | 9.56 | 6.87 | 4.78 | 6.15 | 7.27 | 12.86 |
| Case 20 | 23.6 | 22.6 | 21.9 | 21.2 | 19.9 | 21.6 | 101 | 92 | 101 | 94 | 88 | 90 | 70 | 56 | 69 | 77 | 66 | 62 | 5.33 | 2.74 | 3.18 | 3.79 | 4.21 | 4.99 |

|  | Low Density Lipoprotein (mmol/L) |  |  |  |  |  | Total Cholesterol (mmol/L) |  |  |  |  |  | Triglyceride |  |  |  |  |  | Fatty liver |  |  |  |  |  |
| --- | --- | --- | --- | --- | --- | --- | --- | --- | --- | --- | --- | --- | --- | --- | --- | --- | --- | --- | --- | --- | --- | --- | --- | --- |
|  | Day0 | Day4 | Day8 | Day14 | Day21 | Day51 | Day0 | Day4 | Day8 | Day14 | Day21 | Day51 | Day0 | Day4 | Day8 | Day14 | Day21 | Day51 | Day0 | Day4 | Day8 | Day14 | Day21 | Day51 |
| Case 1 | 4.27 | 5.98 | 6.07 | 5.26 | 4.16 | 3.62 | 7.04 | 7.93 | 8.45 | 7.78 | 6.52 | 6.54 | 2.15 | 1.59 | 1.69 | 1.64 | 1.62 | 2.33 | none | none | none | none | none | none |
| Case 2 | 3.11 | 3.21 | 4.57 | 4.81 | 3.95 | 2.44 | 5.53 | 5.43 | 7.17 | 7.13 | 6.25 | 4.61 | 0.72 | 0.73 | 1.36 | 1.42 | 1.18 | 1.05 | none | none | none | none | none | none |
| Case 3 | 2.25 | 2.82 | 2.96 | 3.18 | 2.90 | 3.27 | 3.80 | 4.51 | 4.48 | 5.00 | 4.43 | 5.54 | 0.94 | 1.10 | 1.08 | 1.30 | 1.24 | 1.39 | none | none | none | none | none | none |
| Case 4 | 3.68 | 5.95 | 7.75 | 6.64 | 4.26 | 3.82 | 6.97 | 8.76 | 10.82 | 9.27 | 7.01 | 6.55 | 3.21 | 2.61 | 1.69 | 1.38 | 1.53 | 2.03 | mi to mo | mi to mo | mild | mild | mild | mild |
| Case 5 | 3.10 | 3.79 | 3.59 | 3.21 | 2.58 | 3.19 | 4.78 | 5.45 | 5.62 | 4.88 | 4.16 | 4.62 | 1.64 | 1.00 | 1.13 | 1.24 | 1.31 | 1.43 | none | none | none | none | none | none |
| Case 6 | 3.93 | 4.76 | 5.66 | 5.05 | 3.86 | 3.73 | 6.39 | 7.36 | 7.96 | 7.65 | 6.13 | 6.27 | 1.19 | 1.56 | 2.18 | 1.99 | 1.53 | 1.28 | none | none | none | none | none | none |
| Case 7 | 3.04 | 5.13 | 6.80 | 4.62 | 3.27 | 1.84 | 5.23 | 7.25 | 8.13 | 6.76 | 4.77 | 3.15 | 2.92 | 2.31 | 2.03 | 2.44 | 2.20 | 1.51 | none | none | none | none | none | none |
| Case 8 | 2.46 | 3.87 | 4.70 | 4.26 | 3.78 | 3.02 | 4.63 | 6.50 | 7.00 | 6.80 | 6.38 | 5.80 | 0.86 | 1.40 | 1.41 | 1.28 | 1.29 | 1.20 | none | none | none | none | none | none |
| Case 9 | 3.52 | 4.55 | 4.58 | 3.93 | 3.20 | 3.22 | 6.11 | 6.27 | 6.49 | 6.11 | 5.21 | 5.60 | 2.22 | 1.40 | 1.37 | 1.30 | 1.38 | 2.03 | moderate | moderate | mi to mo | mild | mild | none |
| Case 10 | 2.66 | 3.14 | 3.21 | 3.24 | 3.25 | 2.55 | 4.32 | 4.95 | 5.22 | 4.96 | 5.37 | 4.20 | 1.04 | 1.52 | 1.07 | 1.10 | 1.33 | 1.10 | severe | severe | mo to se | mo to se | moderate | moderate |
| Case 11 | 3.58 | 4.95 | wd | wd | wd | wd | 5.85 | 6.50 | wd | wd | wd | wd | 1.57 | 1.36 | wd | wd | wd | wd | moderate | moderate | wd | wd | wd | wd |
| Case 12 | 2.82 | 4.10 | 4.64 | 3.28 | 3.15 | wd | 4.59 | 6.53 | 7.01 | 4.82 | 5.54 | wd | 0.87 | 1.04 | 1.07 | 0.93 | 1.08 | wd | none | none | none | none | none | none |
| Case 13 | 4.39 | 5.18 | 5.50 | 5.04 | 4.14 | 4.57 | 6.88 | 7.19 | 7.62 | 7.69 | 6.51 | 7.05 | 1.23 | 1.32 | 1.56 | 1.42 | 1.31 | 1.24 | mi to mo | mi to mo | mi to mo | mild | mild | wd |
| Case 14 | 3.52 | 4.17 | 5.14 | 3.82 | wd | wd | 6.21 | 6.48 | 7.19 | 5.96 | wd | wd | 3.02 | 1.69 | 1.89 | 1.51 | wd | wd | none | none | none | none | wd | wd |
| Case 15 | 4.35 | 5.02 | 5.70 | 4.04 | 3.85 | 3.28 | 6.40 | 7.36 | 8.07 | 6.14 | 6.04 | 5.71 | 1.03 | 0.94 | 1.08 | 1.00 | 1.42 | 1.19 | none | none | none | none | none | none |
| Case 16 | 3.10 | 4.26 | 5.33 | 4.27 | 3.24 | 3.62 | 5.16 | 6.58 | 7.78 | 6.70 | 5.63 | 6.39 | 1.14 | 1.08 | 1.34 | 1.08 | 1.27 | 0.91 | mild | mild | mild | none | none | none |
| Case 17 | 3.35 | 3.75 | 3.58 | 3.28 | 3.23 | 3.28 | 5.24 | 5.69 | 5.47 | 5.36 | 5.13 | 5.22 | 1.54 | 1.63 | 1.32 | 1.36 | 1.33 | 1.22 | severe | severe | mo to se | moderate | mi to mo | mild |
| Case 18 | 4.54 | 4.52 | 3.92 | 3.15 | 2.86 | 4.41 | 7.21 | 7.02 | 6.82 | 5.40 | 5.14 | 7.14 | 1.02 | 0.96 | 1.08 | 1.04 | 0.94 | 1.13 | mild | none | none | none | none | none |
| Case 19 | 2.60 | 3.86 | 4.75 | 5.27 | 4.76 | 4.66 | 4.30 | 6.11 | 6.97 | 7.53 | 7.16 | 7.29 | 1.44 | 2.17 | 1.54 | 1.45 | 1.51 | 2.60 | moderate | moderate | moderate | mi to mo | mild | mild |
| Case 20 | 2.31 | 2.87 | 3.25 | 3.27 | 3.00 | 2.42 | 3.96 | 4.65 | 5.85 | 5.93 | 5.08 | 3.84 | 1.54 | 1.13 | 1.80 | 1.69 | 1.71 | 1.78 | mi to mo | mi to mo | mi to mo | mi to mo | mi to mo | mild |

### Post hoc analysis of health-threatening factors

As abnormal levels of biomarkers leading to increased risk factors of metabolic diseases, we selected clinically relevant cutoffs and compared normal and at-risk subjects for each risk factor (Figure 3), where systolic blood pressure >140mmHg or diastolic blood pressure >90mmHg are associated with hypertension, total cholesterol >6mmol/L and LDL >3.3mmol/L are associated with hyperlipidemia and fasting glucose >6.1mmol/L is associated with hyperglycemia. The data showed that subjects with a systolic blood pressure >140mmHg experienced a greater reduction by 18.6±10.7mmHg in systolic blood pressure than those with normal systolic blood pressure by 3.4±12.4mmHg (P=0.039). However, comparison between participants with diastolic blood pressure >90mmHg and those with normal diastolic blood pressure did not show any difference. Total cholesterol was reduced significantly in participants with baseline levels above 6 mmol/L by 1.3±2.1 mmol/L, while it was slightly elevated in participants with normal baseline levels by 0.76±0.99 mmol/L (P=0.01 between groups). LDL was reduced by 0.34±0.68 mmol/L in those with LDL >3.3 mmol/L but was not reduced in normal-range participants (P=0.016 between groups). Besides, BMI changes between groups with normal range and overweight did not show significant difference (P=0.281). These data indicated that the 21-day fasting had more pronounced effects in subjects with aberrant levels of these health-threatening factors than in subjects with normal levels of these physical indices.

**Figure 3.**
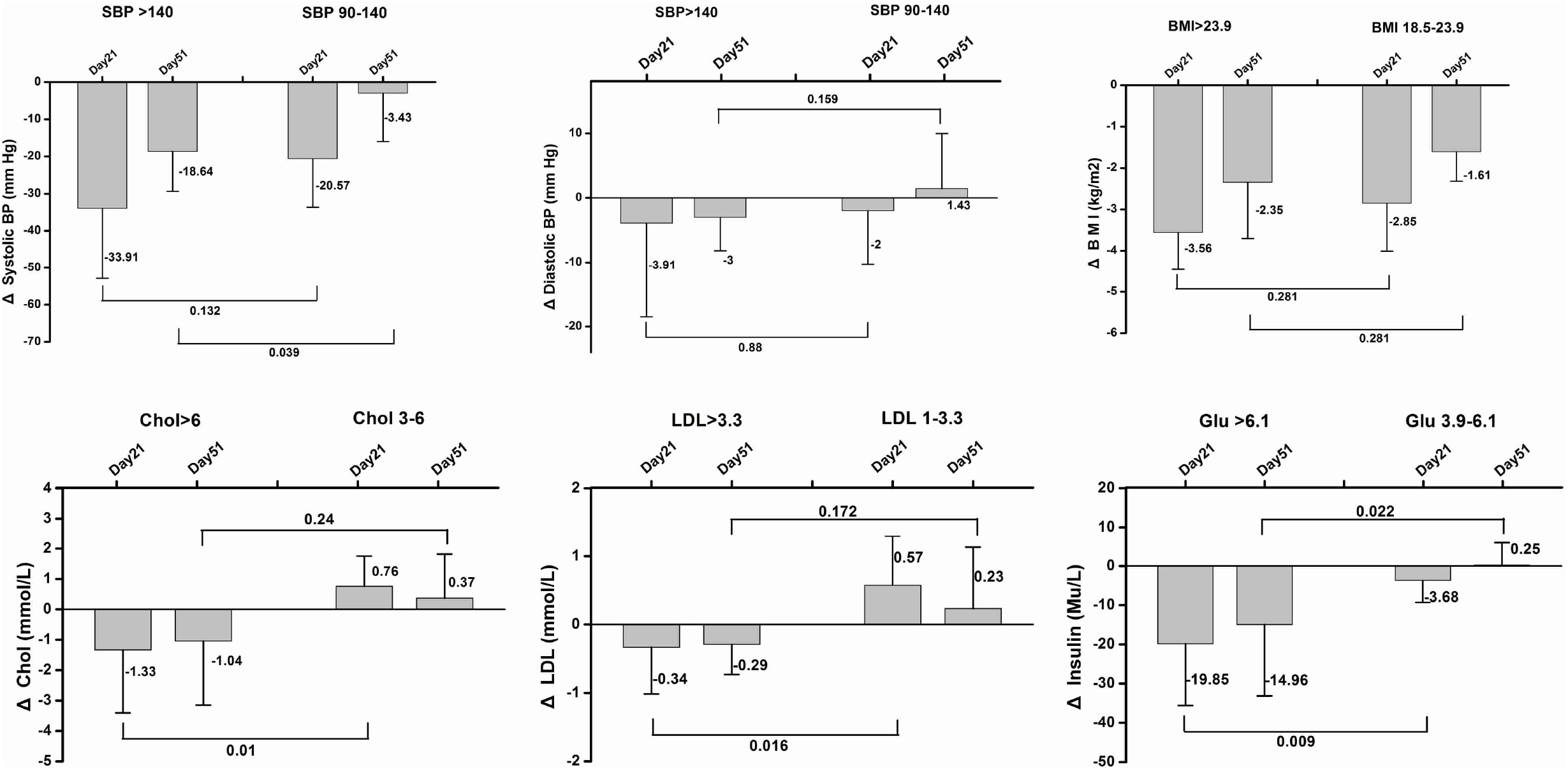
Post hoc analysis of physical biomarkers among subgroups classified by levels of aging/disease risk factors. All the 20 participants were classified into two or three subgroups by the severity of aging/disease risk factors. The Δ value represents a comparison to the baseline. P values were indicated between different subgroups at the same time points.

### Typical case reports

Case #17 was the oldest man among these participants (Table 5). He was a 79 years old and was diagnosed with severe fatty liver just before starting fasting. Then he performed the 21-day fasting with a complete caloric deprivation without any major inconvenience and lost 7.1 kilogram (from 80.6kg to 73.5kg). Levels of his blood pressure, total cholesterol and low-density lipoprotein did not change notably (Figure 4A), while the extent of his fatty liver had a great improvement from severe fatty liver to mild fatty liver on day 51 (Table 5, Figure S1F). Notably, he had not received any treatment for his fatty liver from day 0 to day 51 after starting fasting.

**Figure 4.**
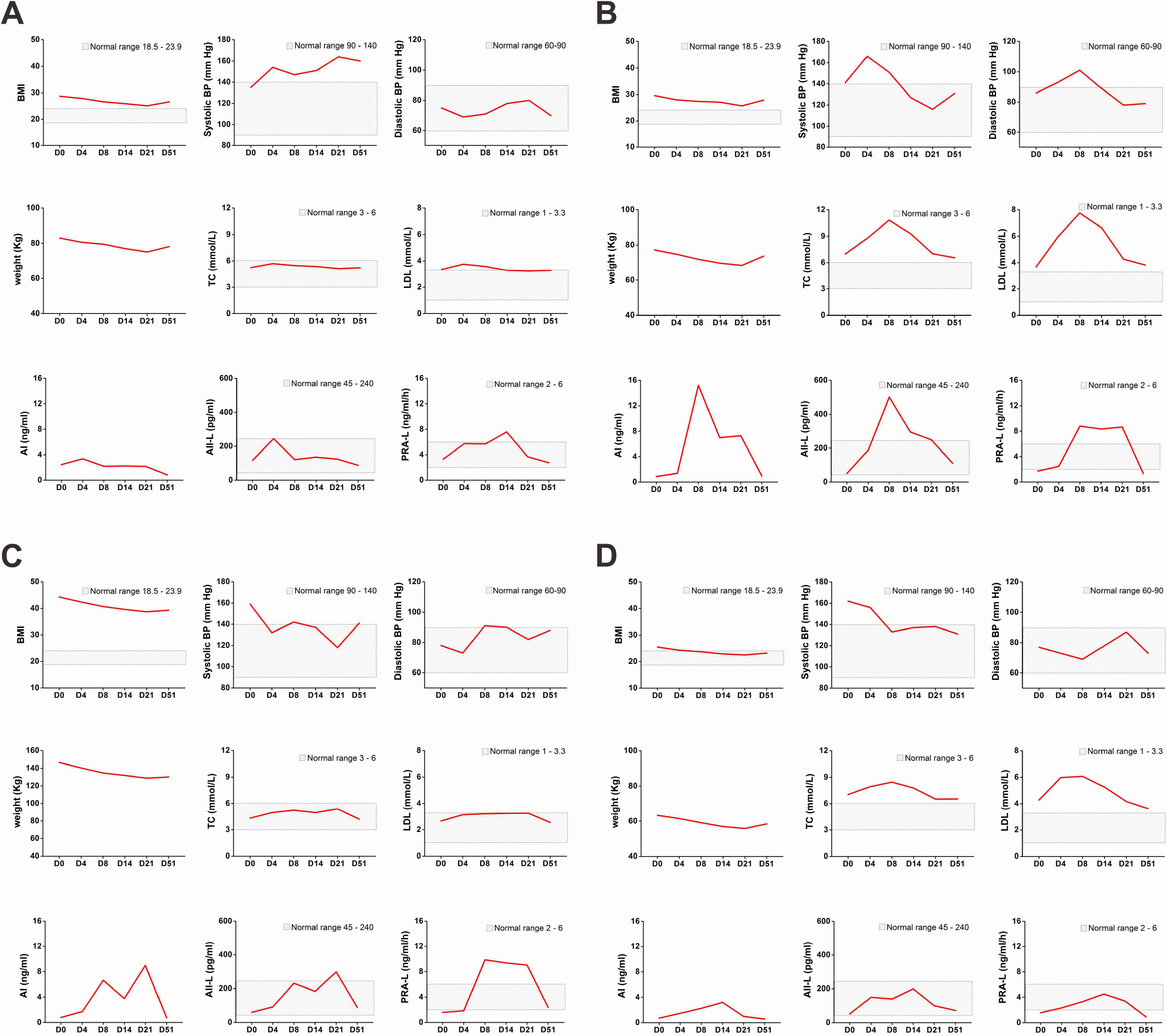
Dynamics of the levels of physical biomarkers for specific cases. The data were examined at indicated time points. Dash areas show the normal ranges.

Case #4 was a 60-year-old Chinese women diagnosed with hypertension and mild-moderate fatty liver (Table 5, Figure S1A). Her BMI was 29.6, much higher than the upper limit value 23.9. Different from other participants, she suffered from varied adverse effects during fasting, including fatigue, weakness, muscle pain, vomiting, headache and sometimes sleep disorder. Her systolic blood pressure also transiently raised from 141mmHg (day 0) to 166mmHg on day 4, but then started to decline and reached to 116mmHg at the end of 21-day fasting (Figure 4B). Her hyperlipemia-associated biomarkers did not change significantly after fasting, though transient increases occurred on day 4, day 7 and day 14 (Figure 4B). After fasting, the extent of her fatty liver turned better from mild-moderate on day 0 to mild on day 51 (Table 5, Figure S1A) through B-sonic without any other treatment.

Case #10 was a 21-year-old Chinese man diagnosed with severe fatty liver, hypertension and overweight. His BMI was 44.3, the highest among all the participants, and his systolic blood pressure was 159 mmHg. He also experienced fatigue, weakness, nausea and vomiting occasionally during fasting. However, his BMI was decreased to 38.7 on day 21 and 39.3 on day 51; and his systolic blood pressure declined to 118mmHg on day 21 and raised to 141mmHg on day 51 (Figure 4C). Meanwhile, the extent of his fatty liver improved from severe on day 0 to moderate on day 51 (Table 5, Figure S1C) without any other treatment.

Case #1 was a 75-year-old Chinese woman diagnosed with hypertension. Before fasting, her BMI was 26.0 and her systolic blood pressure was 162mmHg. After fasting, her BMI was decreased to 22.5 on day 21 and 23.2 on day 51, while her systolic blood pressure declined to 138mmHg on day 21 and 204 on day 51 (Figure 4D). To our biggest surprise, she could still make her stools every 2-3 days even during fasting procedure.

## Discussion

Fasting with different intensities has been performed for a long time out of cultural, religious and medical reasons(*16*). Accumulating evidences show that both calorie restriction and intermittent fasting are effective approaches to minimizing health-threatening factors, ameliorating detrimental inflammations, promoting antitumor therapies as well as extending lifespans(*2, 17*). Recent studies also showed that fasting could modulate immune system by affecting the numbers and functions of monocytes(*18*), B cells(*19*) and T cells(*20*), implicating a therapeutic potential for fasting to treat immune-related diseases(*21*). However, few clinical investigations on the effects of long-term (> 7 days) fasting have been conducted due to either the poor subject compliance or lack of systemic scientific observation and records. In this study, a well-organized Bigu camp with participants of good obedience has been followed in a timely and scientifically way, by which time serial dynamic changes of their physical indices could be recorded.

From our preliminary investigation, we found that at the 21-day fasting with <5% calorie intake was applicable for normal people. No severe adverse effects greater than grade 2 were reported. Physical examination data also suggested that nearly all biological markers fell within the safe limits during fasting, except for the uric acid, which appeared beyond the normal range for nearly all particants. This should be caused by inhibition of renal excretion of uric acid by the ketosis of fasting(*22*). During the whole fasting procedure, either hypoglycemia or hyponatremia or hypokalemia has not been observed. Therefore, it is presented that the long-term extreme fasting following Bigu regimen seemed very likely to be safe for both healthy and un-healthy people.

Our data also showed that this long-time extreme fasting could help reduce health-threatening factors for participants with hypertension and hyperlipidemia. The data suggested that after fasting, the levels of BMI, diastolic blood pressure, systolic blood pressure, blood urea nitrogen, total cholesterol, low density lipoprotein and triglyceride were significantly decreased among all the participants. Compared to the previously reported clinical trials using fasting-mimicking diet(*2*), which presented significant decreases of the levels of BMI, diastolic blood pressure and systolic blood pressure, and decrease but not significant of the levels of total cholesterol, low density protein and triglyceride, our data showed all markers were significantly decreased. Moreover, all the above indices showed excessive reductions following our fasting procedure than in the reported clinical trials. Since the long-term extreme fasting could result in more drastic decreases of cardiovascular associated biomarkers, it would lead to a better improvement of health.

Nonalcoholic fatty liver disease (NAFLD) is one of the most common liver diseases in parallel with obesity and other metabolic diseases (*23, 24*). It is driven by an access accumulation of metabolic substrates, e.g. fatty acid(*25, 26*), and is considered to be a high risk factor of hepatocellular carcinoma(*27*). However, its therapeutic approaches are still limited, except for changing of life styles, especially diet (*23, 28*). Scientists found that the Mediterranean diet, with less intake of fatty acid, demonstrated to be of high benefit for NAFLD patients(*29*). In our study, a 21-day extreme fasting period could help minimize the intake of fatty acid but keep normal activities. More importantly, of the 9 participants with fatty liver, 7 were ameliorated, including 2 from severe to moderate or mild, 4 from moderate to mild, and 1 from mild to non-fatty liver (Figure S1A-I). These data suggested that long-time extreme fasting could be exploited as a safe and effective therapeutic approach for NAFLD patients.

In summary, we investigated the traditional Chinese long-time extreme fasting (Bigu regimen) using the evaluation system of modern medical sciences for the first time. The long-term extreme fasting appeared not only safe and feasible for both healthy and unhealthy people, but also could ameliorate the risk factors associated with hypertension and hyperlipidemia. Our study encouraged the further investigations on fasting and its therapeutic effects.

## Data Availability

All the data have already been attached in supplementary dataset and the raw data can be accessible once requested.

## Acknowledgments

We thank Mr. Feng Liu, Mingjun Han and Yuanlei Chen for their help in the organization of this clinical trial and Beijing Tianyi Medical Technology Limited Company for their help in sample collection.

## Author Contributors

MZ, DL, HD, CW and LM designed the study. CW, LM, QW, TC, LW, ZZ, DT, WL and YW analysed the data. CW, LM, LJ, QW, TC, LW, HD, DL and MZ interpreted the data. CW, LM, QW, TC and DL prepared the figures. CW, QW, DL and MZ wrote the paper. All authors approved the final manuscript as submitted and agree to be accountable for all aspects of the work.

